# Impact of COVID-19 on clinical practice, income, health and lifestyle behavior of Brazilian urologists

**DOI:** 10.1101/2020.06.05.20121566

**Authors:** Cristiano M. Gomes, Luciano Favorito, João Victor T. Henriques, Alfredo F. Canalini, Karin M. J. Anzolch, Roni de C. Fernandes, Carlos H S Bellucci, Caroline S Silva, Marcelo L. Wroclawski, Antonio Carlos L. Pompeo, Jose de Bessa

## Abstract

**Objectives:** To evaluate the impact of COVID-19 on clinical practice, income, health and lifestyle behavior of Brazilian urologists during the month of April 2020.

**Materials and Methods:** A 39-question, web-based survey was sent to all urologist members of the Brazilian Society of Urology. We assessed socio-demographic, professional, health and behavior parameters. The primary goal was to evaluate changes in urologists’ clinical practice and income after two months of COVID-19. We also looked at geographical differences based on the incidence rates of COVID-19 in different states.

**Results:** Among 766 urologists who completed the survey, a reduction ≥ 50% of patient visits, elective and emergency surgeries was reported by 83.2%, 89.6% and 54.8%, respectively. An income reduction of ≥ 50% was reported by 54.3%. Measures to reduce costs were implemented by most. Video consultations were performed by 38.7%. Modifications in health and lifestyle included weight gain (32.9%), reduced physical activity (60.0%), increased alcoholic intake (39.9%) and reduced sexual activity (34.9%). Finally, 13.5% of Brazilian urologists were infected with SARS-CoV-2 and about one third required hospitalization. Urologists from the highest COVID-19 incidence states were at a higher risk to have a reduction of patient visits and non-essential surgeries (OR = 2.95, 95% CI 1.86 – 4.75; p < 0.0001) and of being infected with SARS-CoV-2 (OR = 4.36 95%CI 1.74–10.54, p = 0.012).

**Conclusions:** COVID-19 produced massive disturbances in Brazilian urologists’ practice, with major reductions in patient visits and surgical procedures. Distressing consequences were also observed on physicians’ income, health and personal lives. These findings are probably applicable to other medical specialties.

## 1. Introduction

The COVID-19 pandemic has had broad consequences beyond the spread of the disease itself and efforts to restrain it. A major aspect of the pandemic is the fact that it caused the most extensive recession in history, with more than a third of the global population at the time being placed on lockdown.(1, 2)

As we prepare this manuscript, Brazil’s curve of new cases and deaths is probably at its peak.(3) Social distancing has been introduced for more than two months and is plainly effective throughout the country, with no signs that it will be relaxed in the next weeks.(2)

The pandemic has disrupted daily professional activities and personal life routine.(4) For medical professionals, regular patient care was dramatically affected. The focus of medical care for both the public and private health systems shifted to the management of patients with Covid-19 and avoidance of any non-emergency treatments.(5)

In urology, surgery is limited to emergencies.(6–8) Elective surgeries have been suspended and even cancer cases have been postponed, with the hope that such delay will not have a negative impact on patients’ outcome.(9) Telemedicine has become an important tool, reducing the exposure of patients and clinicians, but only a small fragment of practitioners was prepared for this and most patients do not openly accept this new way modality of evaluation.(10, 11) As a result, urologists and other surgical specialists have been experiencing a tremendous impact on their income, especially those whose earnings are driven mostly from their private practice.(12) As other medical specialists and dentists, most costs with personnel and rent have been maintained in spite of their dramatically reduced income. Many are facing a financial crisis which is expected to worsen.

Apart from the economic and professional impacts, COVID-19 is attacking people’s personal lives. The unprecedented restrictions to movement and the isolation of those under greater risk have created unthinkable situations and forced major changes in lifestyle.(2) Big changes in everyday routines due to the need to stay at home and the widespread closures of schools along with the financial hardships and fear of losing a loved one to the disease may raise the level of anxiety and tension in family relationships.(13, 14)

The aim of this study is to performed a cross-sectional evaluation of the impact of COVID-19 on clinical practice, income, health and lifestyle behavior of Brazilian urologists.

## 2. Materials and Methods

This study was approved by the Research Ethics Committee of the University of Sao Paulo School of Medicine (project number CAPPESQ 13051/2020) and informed consent was obtained from all participants.

This study was conducted as an electronic survey sent through e-mail to all urologist members of the Brazilian Society of Urology, with no incentives for completion. The first email inviting urologists to participate was sent on April/29/2020; two additional invitations were sent on May/04 and May/06/2020. Data collection was closed on May/10/2020. The data reported in this study refers to urologists’ activities and impressions during the month of April/2020, when the whole country was living the second month of social distance officially recommended by state and federal sanitary authorities.

The invitation to participate was sent to 4200 eligible urologists. The invitation e-mail contained a link to a 39-question, web-based survey (supplementary file 1). Most questions were closed-ended, multiple choice. Some also allowed an open answer.

The survey included an assessment of socio-demographic, professional, health-related and behavior parameters. The primary goal was to collect data on changes in urologists’ clinical practice and income after 2 months of COVID-19. We also investigated changes in urological indications and planning for the near future regarding cost containment in professional and personal life. Urologists were asked to estimate the time beyond which delaying a surgery would be harmful to patients based on a priority tier classification varying from 1 to 3 that was recommended by the European Association of Urology (EAU).(15)

We also investigated the impact of COVID-19 on health parameters and lifestyle behaviors.

Given the fact that Brazil is a country with continental dimensions and the incidence of COVID-19 varies markedly between different states, we investigated the impact of COVID-19 incidence on urologists’ practice and income. Based on official data provided by the Brazilian Health Minister(3), we compared data from the 3 states with more participants in this survey that were among the lowest incidence rates states (Minas Gerais, Rio Grande do Sul and Santa Catarina) with the 3 states with more participants among the highest incidence states (São Paulo, Rio de Janeiro and Pernambuco).

### Statistical methods

Data were initially elaborated using Survey Monkey® software online. Quantitative variables were expressed as medians and interquartile ranges, while qualitative variables were expressed as absolute values, percentages, or proportions. categorical variables were compared using the Chi-squared or Fisher’s exact test. Association were described as Odds Ratio with respective confidence intervals. All tests were 2-sided and a p value < 0.05 was considered statistically significant. GraphPad Prism, version 8.0.4, San Diego-CA, USA, was used for data analysis

## 3. Results

A total of 766 subjects completed the survey, representing 18.2% of the invited urologists. The mean time to complete the questionnaire was 12 minutes and 57 seconds. The skipping rate for each question ranged from 0% (Q1) to 38.6% (Q39). Most responses (83.4%) were filled during 4 of the 12 days that the survey was open.

The median age of the participants was 46.0 years [38–57]. Most were (90.8%), married (92.2%) and have children 80.4%. The state and geographic region distribution of the participants are shown in Table 1 as well as the actual distribution of Brazilian urologists through states and geographic regions. The participation of urologists from the five different Brazilian geographic regions was proportional to the actual distribution of Brazilian urologists (p = 0.881).

**Table 1.** Geographic regions in Brazil and state distribution of the 766 participants of the survey

| Geographic region / State | N* (%) | Actual state distribution** (%) |
| --- | --- | --- |
| Southeast | 361 (49.9) | 55.5 |
| Sao Paulo | 231 (31.9) | 31.5 |
| Minas Gerais | 54 (7.5) | 11.0 |
| Rio de Janeiro | 59 (8.2) | 11.0 |
| Espirito Santo | 17 (2.3) | 2.0 |
| South | 141 (19.3) | 15.4 |
| Parana | 32 (4.4) | 5.6 |
| Rio Grande do Sul | 65 (8.9) | 6.1 |
| Santa Catarina | 44 (6.0) | 3.7 |
| Northeast | 144 (19.8) | 16.7 |
| Alagoas | 10 (1.4) | 0.6 |
| Bahia | 38 (5.2) | 4.7 |
| Ceara | 18 (2.5) | 2.9 |
| Maranhao | 8 (1.1) | 1.2 |
| Paraiba | 6 (0.8) | 1.1 |
| Pernambuco | 35 (4.8) | 3.3 |
| Piaui | 11 (1.5) | 0.9 |
| Rio Grande do Norte | 11 (1.5) | 1.4 |
| Sergipe | 7 (1.0) | 0.6 |
| Center-West | 53 (7.3) | 8.5 |
| Federal District | 15 (2.0) | 2.9 |
| Goias | 24 (3.3) | 3.2 |
| Mato Grosso | 7 (1.0) | 1.1 |
| Mato Grosso do Sul | 7 (1.0) | 1.3 |
| North | 26 (3.7) | 3.7 |
| Acre | 0 (0.0) | 0.1 |
| Amapa | 1 (0.2) | 0.2 |
| Amazonas | 6 (0.8) | 0.7 |
| Para | 15 (2.0) | 1.4 |
| Rondonia | 1 (0.2) | 0.6 |
| Roraima | 1 (0.2) | 0.2 |
| Tocantins | 2 (0.3) | 0.5 |

\* Urologists who participated in the survey
\*\*Official state distribution of urologists who received invitation for the survey

The distribution of urologists according to the type of medical practice is depicted in figure 1. The majority (61.2%) has most of their earnings coming from work in private practice, followed by employment in the public health system (14.6%) and employment in the private sector (14.1%). These numbers vary by region, with some states having a major preponderance of independent practice while others being dominated by the employed physician model.

**Figure 1.**
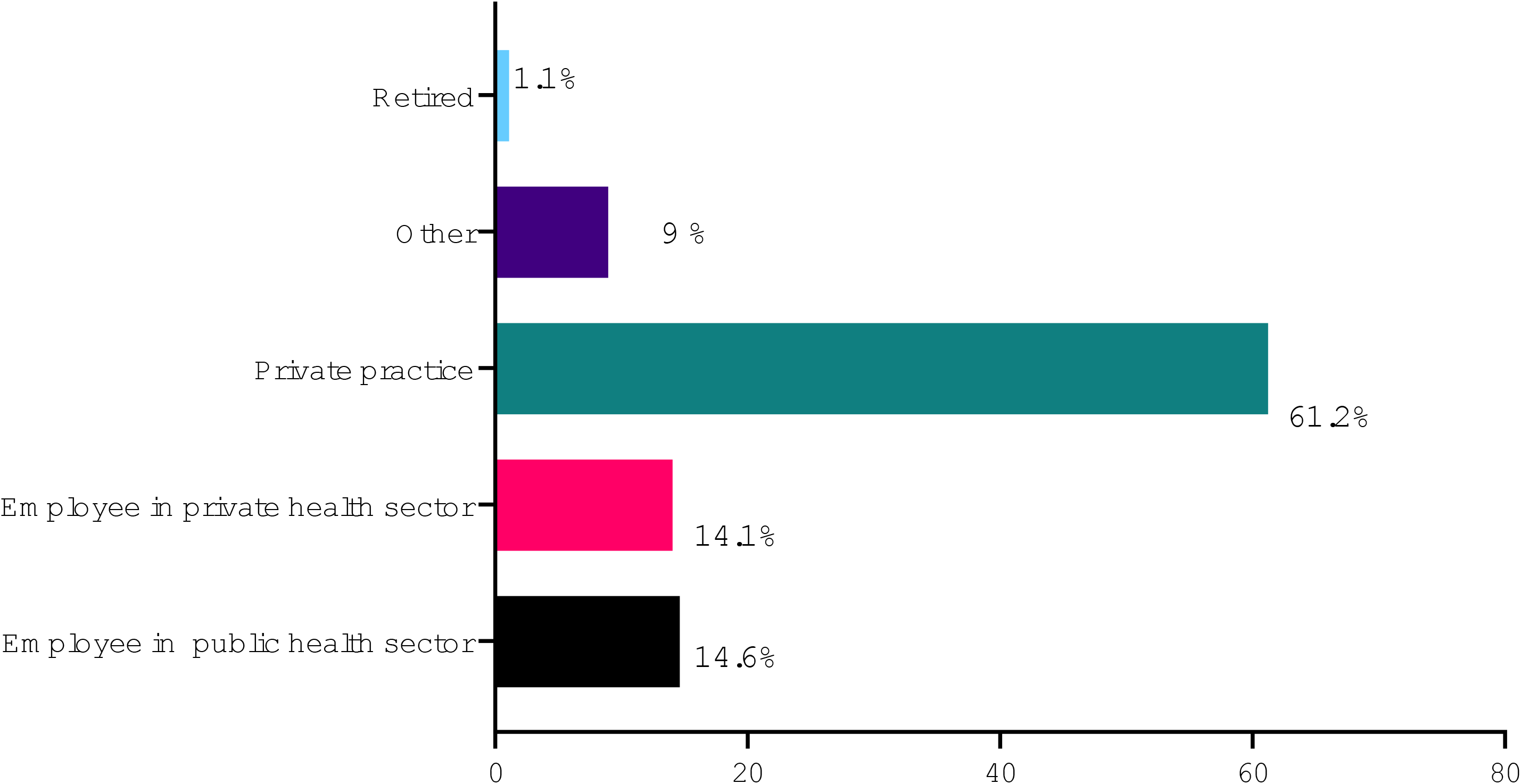
Distribution of urologists according to type of medical practice

The enormous impact of COVID-19 on urologists’ clinical practice is shown in Table 2, with a major reduction in daily hours of work, patient visits, elective and emergency surgeries.

**Table 2.** Impact of COVID-19 on Brazilian urologists’ clinical practice

| Practice activity | N (%) |
| --- | --- |
| Daily hours worked on weekdays |  |
| None | 79 (10.9) |
| Up to 2 hours/day | 140 (19.3) |
| Between 2-4 hours/day | 217 (29.9) |
| Between 4-8 hours/day | 225 (31.0) |
| > 8 hours/day | 64 (8.9) |
| Elective patient visits |  |
| Remained stable | 3 (0.4) |
| Decreased up to 25% | 23 (3.2) |
| Decreased 25 to 50% | 96 (13.2) |
| Decreased 50 to 75% | 205 (28.3) |
| Decreased > 75% | 398 (54.9) |
| Elective surgeries |  |
| Remained stable | 5 (0.7) |
| Decreased up to 25% | 14 (1.9) |
| Decreased 25 to 50% | 50 (6.9) |
| Decreased 50 to 75% | 98 (13.5) |
| Decreased > 75% | 551 (76.1) |
| Increased | 1 (0.2) |
| Emergency surgeries |  |
| Remained stable | 116 (16.0) |
| Decreased up to 25% | 91 (12.6) |
| Decreased 25 to 50% | 108 (14.9) |
| Decreased 50 to 75% | 141 (19.4) |
| Decreased > 75% | 256 (35.4) |
| Increased | 7 (3.3) |

According to the surgical priority classification recommended by EAU, emergency surgeries were performed by 79.6% of the participants, priority 1 surgeries by 51.3%, priority 2 by 34.1% and priority 3 by 47.0%, in April/2020. Non-essential procedures were performed by 31.3% of the participants during this period.

Urologists’ estimates of time beyond which delaying surgeries may be harmful to patients are shown in Table 3. Most urologists considered that delaying surgeries classified as Priority 1 for more than 1 month might be harmful for the patient. Delays of up to two months for Priority 2 and up to 3 months for Priority 3 surgeries were considered acceptable by most urologists.

**Table 3.**
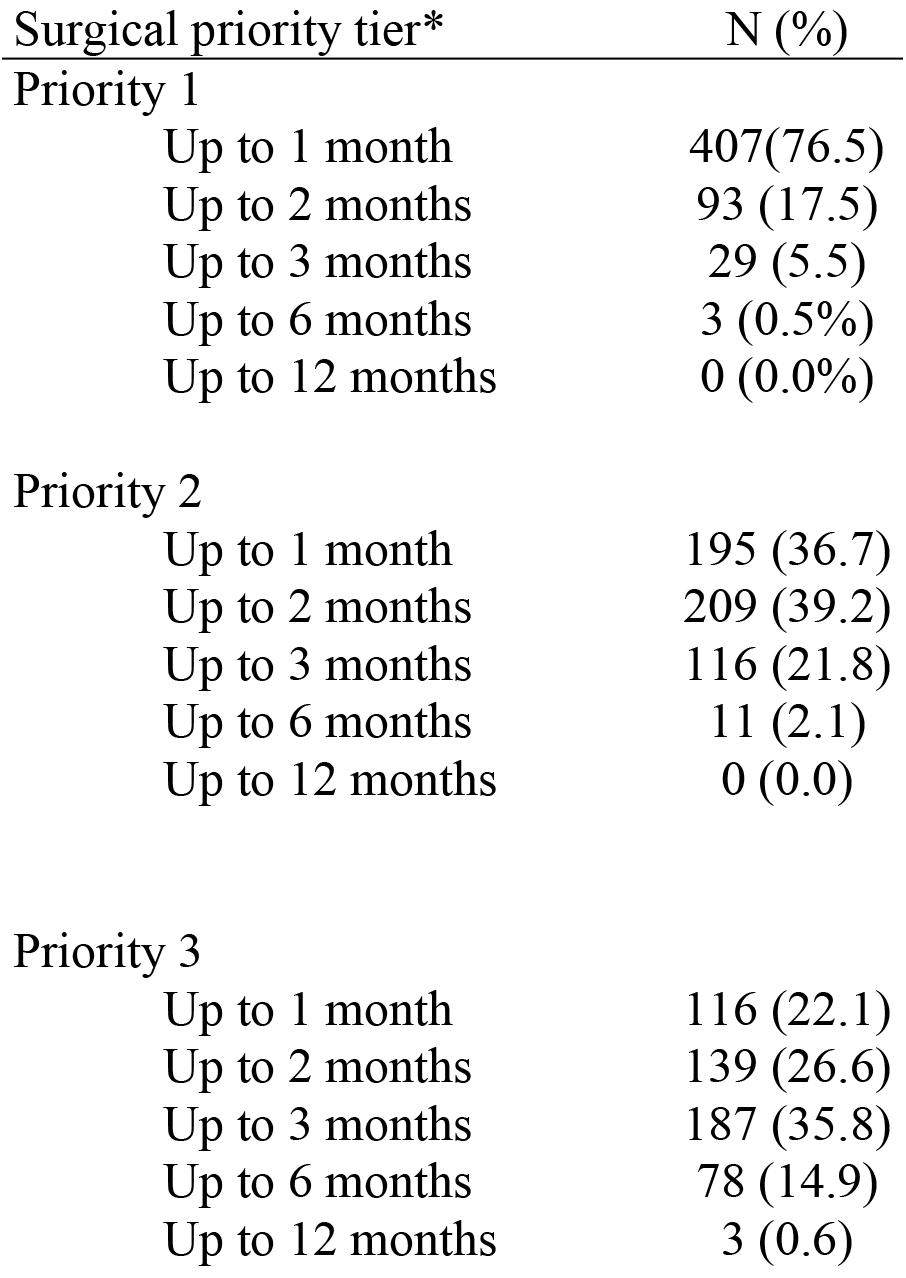
Urologists’ estimates of time beyond which delaying surgeries would harm patients

The impact of COVID-19 on urologists’ income is shown in Figure 2. An income reduction of ≥ 50% was reported by 54.3% of the participants, including 23.8% who had a > 75% income reduction. Urologists in private practice had a higher chance of experimenting a > 75% income reduction in comparison with those working as employees in private or public sector (OR = 3.69, 95% CI 2.09–6.35; p < 0.0001).

**Figure 2.**
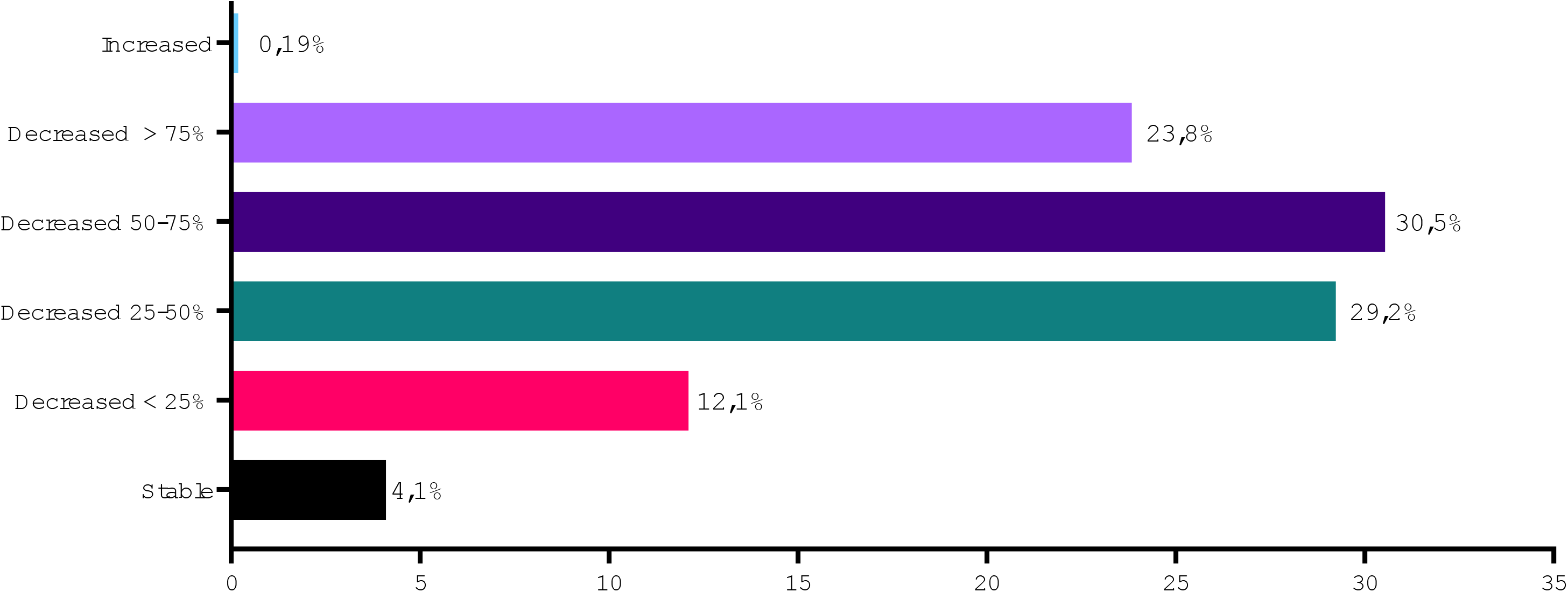
Impact of COVID-19 pandemic on urologists’ income

Since the start of the COVID-19 pandemic, 38.7% of the urologists reported performing video consultations. Patients’ acceptance of video consultations was high as was physician’s confidence regarding proper diagnosis and management using this consulting modality. A fee for service was charged in more than 50% of video consultations. (Table 4)

**Table 4.** Patient acceptance, physician’s confidence and charging for video consultations during the COVID-19

| Video consultations | N (%) |
| --- | --- |
| When you proposed video consultations, what percentage of patients accepted? |  |
| > 80% of the patients | 87 (41.6) |
| between 50-80% of the patients | 44 (21.1) |
| < 50% of the patients | 78 (37.3) |
| How confident were you with the diagnosis and treatment of patients during video consultations? |  |
| Confident in > 80% of consultations | 103 (49.2) |
| Confident in 50 to 80% of consultations | 62 (29.7) |
| Confident in < 50% of consultations | 44 (21.1) |
| When you performed video consultations, how often you charged a fee? |  |
| > 80% of consultations | 103 (49.3) |
| 50 to 80% of consultations | 24 (11.5) |
| < 50% of consultations | 82 (39.3) |

Urologists’ measures to reduce costs in professional practice and personal life since the start of COVID-19 are described in Table 5.

**Table 5.** Reduction of costs in professional practice and personal life since start of COVID-19

| Measures to reduce costs | N (%) |
| --- | --- |
| Professional practice |  |
| None | 271 (51.6) |
| Fired one or more an employee | 102 (19.4) |
| Reduced investment in advertising/marketing | 141 (26.9) |
| Returned rented office | 12 (2.3) |
| Closed office | 33 (6.3) |
| Other | 57 (10.8) |
| Personal life |  |
| None | 158 (30.1) |
| Children's education | 43 (8.2) |
| Healthcare plan | 14 (2.7) |
| Supermarket | 181 (34.5) |
| Housekeeper | 133 (25.3) |
| Phone, internet, TV | 83 (15.8) |
| Other health professionals (psychotherapist, dentist) | 115 (21.9) |
| Gym/personal trainer | 237 (45.1) |
| Returned rented vacation home | 6 (1.1) |
| Sold car, motorcycle, boat | 10 (1.9) |
| Music, dance, theater or similar classes | 121 (23.0) |
| Foreign language classes | 48 (9.1) |
| Other | 48 (9.1) |

Table 6 compares the impact of COVID-19 on urologists’ clinical practice from states with highest vs lowest COVID-19 incidence. Reduction of elective patient visits were more severely reduced in states with highest incidence of COVID-19. Almost 90% of the urologists from states with highest incidence of COVID-19 reported > 50% reduction of elective patient visits vs 70% of the participants from states with low incidence of COVID-19 (OR = 2.95, 95% CI 1.86 – 4.75; p < 0.0001). The impact on daily hours worked, emergency and elective essential surgeries (Priority 1 to 3 surgeries) was not significantly different based on the incidence of COVID-19. The chance of performing non-essential procedures was lower among urologists living in the highest incidence COVID-19 states (OR = 0.52 95% CI 0.33–0.83, p = 0.009).

**Table 6.** Urology practice in states with highest vs lowest incidence of COVID-19

| Urology practice parameters | Highest incidence states | Lowest incidence states | OR (95% CI) | *p* |
| --- | --- | --- | --- | --- |
| Daily hours worked |  |  | 1.17 (0.80-1.71) | 0.438 |
| Decreased < 4 hours | 61.0% | 57.0% |  |  |
| Decreased > 4 hours | 39.0% | 43.0% |  |  |
| Elective patient visits |  |  | 2.95 (1.86-4.75) | < 0.001 |
| Decreased < 50% | 12.4% | 29.4% |  |  |
| Decreased > 50% | 87.6% | 70.6% |  |  |
| Emergency surgeries |  |  | 0.98 (0.66-1.43) | >0.999 |
| Decreased < 50% | 44.8% | 44.4% |  |  |
| Decreased > 50% | 55.2% | 55.6% |  |  |
| Elective essential surgeries* |  |  | 1.40 (0.76-2.57) | 0.268 |
| Decreased < 50% | 9.1% | 12.4% |  |  |
| Decreased > 50% | 90.9% | 87.6% |  |  |
| Non-essential surgeries** |  |  | 0.52 (0.33-0.83) | 0.009 |
| At least one surgery | 24.5% | 38.4% |  |  |
| No surgeries | 75.5% | 61.6% |  |  |

\*Priority 1 to 3 surgeries; \*\* percentage of surgeons who performed at least one non-essential surgery

The impact on income was profound and similar between states with highest vs lowest incidence of COVID-19. Among urologists living in highest incidence COVID-19 states, 23.6% reported an income reduction > 75% as opposed to 21.5% in those from lowest incidence states (OR = 1.13, 95% CI 0.67–1.94; p = 0.692).

Asked about their physical health, 357 (68.0%), 140 (26.7%) and 28 (5.3%) considered themselves as very healthy, moderately healthy and not healthy, respectively. The median body mass index was 26.5 Kg/cm^2^ [24.4 – 28.7]. The impact of COVID-19 on urologists’ weight, physical activities, alcohol consumption and sexual activity is shown in table 7.

**Table 7.** Changes in weight, physical activity, alcohol consumption and sexual activity during COVID-19

| Health parameters during COVID-19 | N (%) |
| --- | --- |
| Weight | N (%) |
| Slightly reduced (less than 3Kg) | 80 (15.2) |
| Significantly reduced (more than 3Kg) | 22 (4.2) |
| Stable | 236 (44.9) |
| Slightly increased (less than 3 Kg) | 140 (26.6) |
| Significantly increased (more than 3Kg) | 33 (6.3) |
| I don't know | 14 (2.7) |
| Physical activity |  |
| Reduced | 315 (60.0) |
| Stable | 124 (23.6) |
| Increased | 86 (16.4) |
| Alcoholic beverages intake* |  |
| Reduced | 88 (21.6) |
| Stable | 157 (38.5) |
| Increased | 163 (39.9) |
| Sexual activity |  |
| Reduced | 164 (34.9) |
| Stable | 243 (51.7) |
| Increased | 63 (13.4) |

\*participants who reported not drinking alcoholic beverages were removed from calculations

The activities that occupied most of the urologists’ time out of working hours during the two first months of the COVID-19 pandemic were online social media, housekeeping, watching movies and TV shows and spending time with family (Table 8).

**Table 8.**
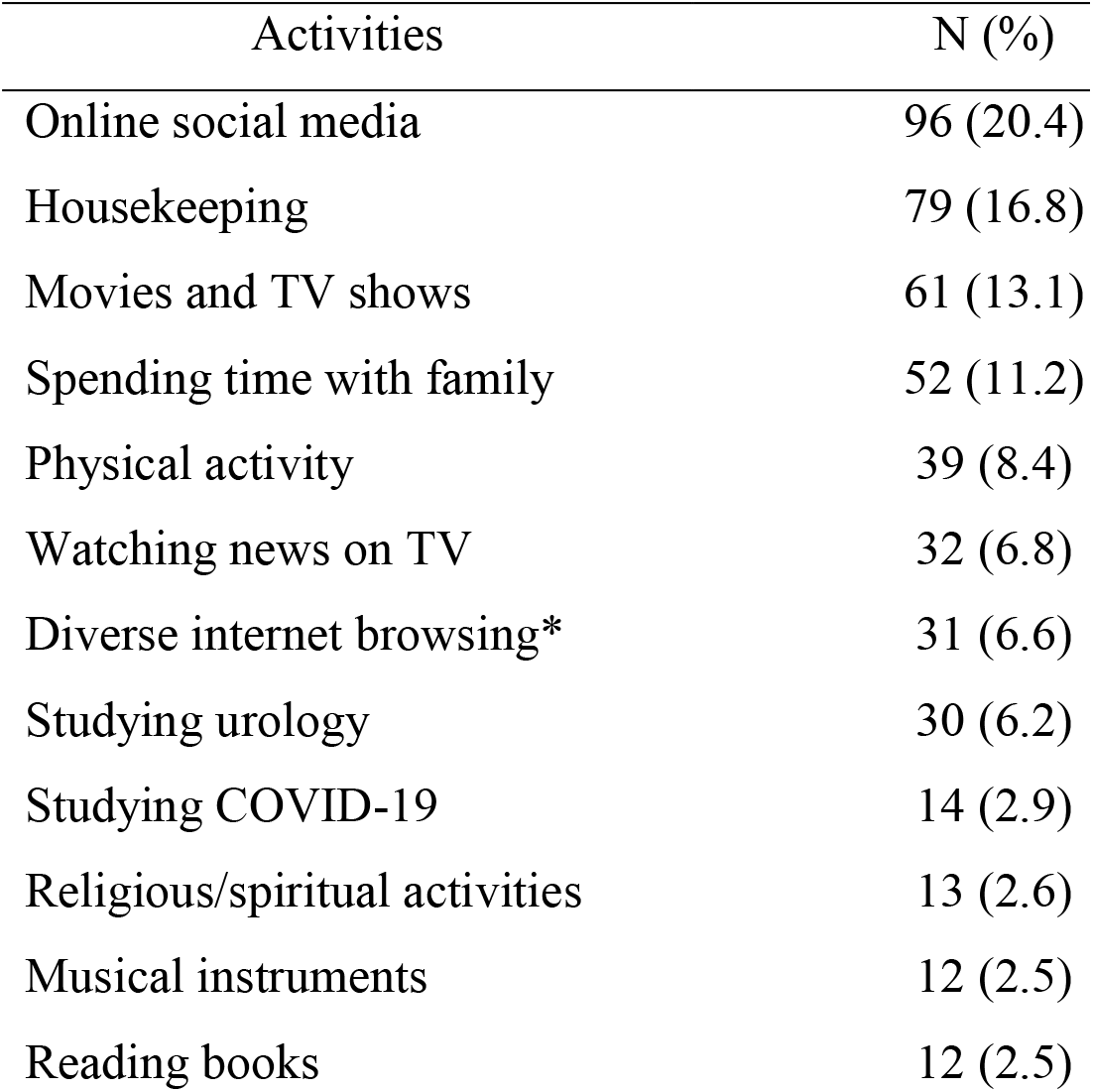
Activities on non-working hours during COVID-19 pandemic

At the end of April/2020, after two months of the pandemic in Brazil, 13.5% of Brazilian urologists have had COVID-19, including 5.2% with unequivocal laboratory confirmation and 8.2% with diagnosis based on clinical, radiological and epidemiological parameters. In most instances the clinical presentation was mild, but 34.8% of the affected urologists required hospitalization. Urologists living in the highest incidence COVID-19 states had much higher chance of having COVID-19 than those living in lowest incidence states (17.1% vs 4.5%; OR = 4.36 95%CI 1.74–10.54, p = 0.012).

## 4. Discussion

The coronavirus pandemic is a major challenge for healthcare systems worldwide and has impacted the habits, economy and health systems around the world. The routine of all urologic procedures has changed and, in Brazil, urology practice was drastically affected.(16, 17) As we prepare this manuscript, in late May/2020, social distancing remains fully effective in most areas in Brazil, with no clear perspective about when it will be relaxed.(2, 3)

In this study we have shown several and striking changes affecting Brazilian urologists during the first two months of COVID-19 pandemic. Urologists have had a major reduction of patient visits, elective and emergency surgeries. Non-essential surgeries have been performed by a minority of Brazilian urologists since the outbreak of COVID-19. Most have experienced a tremendous cutback in their income, especially those in private practice, prompting measures to reduce costs in professional practice and personal life. Along with such major professional changes, significant modifications in health and lifestyle have also been observed, including weight gain, reduced physical activity, increased alcoholic beverages intake and reduced sexual activity. Finally, 13.5% of participant urologists were infected with SARS-CoV-2 and about one third of them required hospitalization. As we prepare this manuscript, COVID-19 had taken the lives of 4 Brazilian urologists, to whom we dedicate this study.

A total of 4200 urologists received a link to the interview and 766 (18.2%) participated. The distribution of respondents was in accordance with the actual distribution of urologists throughout the different states and geographical areas of the country, confirming a well-balanced and representative participant distribution. The skipping rate for each question was satisfactory, ranging from 0% to 38.6% and the mean time to completion was almost 13 minutes. Because we gave no incentives to participants and we used a long questionnaire (39 questions), we consider our participation rate of 18.2% as very good, and in line with recent online surveys evaluating practice patterns by healthcare providers in which participation rates varied from 2% to 25%.(18–22)

The outbreak of COVID-19 has drastically changed how outpatient care is delivered in health care practices. To decrease the risk of contamination for patients and health care workers, providers are deferring elective and preventive visits as well as non-emergency surgeries. For their part, patients are also avoiding them to prevent unnecessary exposure. Also influencing these behaviors are the authorities’ recommendations to restrict travel and nonessential services. Many national health authorities are recommending cancellation of scheduled elective surgeries and office visits.(15, 23, 24) The objectives are to increase the availability of inpatient and intensive care units’ beds, to permit internal redeployment of medical staff and nurses, and to avoid overwhelming the health care system. Some reports have pointed that the number of visits to ambulatory care practices declined by nearly 60 percent in the pandemics. (5)

Our results are in accordance with this context, showing a profound impact on Brazilian urologists’ clinical practice, with a drastic reduction of the number of patient visits and all types of surgical cases. A reduction ≥ 50% of elective surgeries was reported by 80% of the participants. Interestingly, a decline in emergency surgeries was also reported in large scale. Only 16% reported that emergency surgeries remained stable while more than 50% revealed a ≥ 50% decrease of these procedures. Consequently, a tremendous reduction of hours worked per day was observed, with 30.2% of the urologists working less than 2 hours per day and only 8.9% working more than 8 hours per day during the month of April/2020.

Since Brazil is a country with continental dimensions and the incidence of COVID-19 varies markedly between different states, we investigated the impact of COVID-19 incidence by comparing data from 3 states among those with the lowest incidence rates with 3 states among the highest incidence states.(2, 3) We found that urologists from the highest COVID-19 incidence states were at a higher risk to have a major reduction of patient visits and elective surgeries. The most likely explanation for this is the earlier and more vigorous implementation of social distancing measures, with reduced mobility. A greater fear of contamination in these areas may possibly make urologists and patients more inclined to postpone elective consultations and surgeries.

The effect of the observed changes in urology practice on income was also of enormous proportions, with more than 95% of the urologists reporting a significant reduction in their earnings, including 54.3% with a decline of ≥ 50% during this period. Urologists in private practice (61.2% of the participants) were more profoundly affected and had a 3.7 times higher chance of experimenting a > 75% income reduction in comparison with those working as employees in the private or public sector. Such major reduction in practice and income has been reported in other countries and with other medical specialties.(5, 12, 25–29) Consistent with the observed income cutback, a great proportion of Brazilian urologists have already implemented cost containment measures in their professional and personal lives.

Because of the pandemics, remote care through telemedicine has been recently regulated and temporarily authorized by the Brazilian Ministry of Health.(30) Before COVID-19, telemedicine was not in practice by the vast majority of Brazilian physicians and was restricted to specific projects. The fact that almost 40% of Brazilian urologists have had experience with video consultations is such short period is remarkable. Another significant finding was that urologists were reimbursed in most video consultations. We did not evaluate how this was done and how much was charged. We also did not evaluate differences in how telemedicine has been used in the private and public health systems. Because telemedicine has only recently been introduced in Brazil, billing regulations are not well stablished yet.

Our study also showed that most physicians were confident with their diagnosis and treatment upon the use of video consultations and most patients seem to accept it and be content with it. Indeed, studies have shown high levels of effectiveness and patient satisfaction with tele-interventions in many clinical scenarios.(31, 32) Telemedicine limitations, however, must be discussed constantly with the medical and lay population. We believe that tele-screening, test reviews and follow-up evaluations that do not require physical examination are the ideal situations for this type of care, especially when the patient is in the high-risk group and must be socially isolated.(33)

During the pandemics, the general recommendation is to reschedule surgical procedures that can be postponed without harm to the patient, reducing the risks of contamination for the patient and the surgical team. It is not well defined which operations can be postponed and for how long. Different guidelines and recommendations have been published in order to help practitioners and authorities in prioritizing essential surgeries.(15, 17) These recommendations, however, are mostly not evidence-informed, but based on expert opinion. Most urologists in Brazil did not perform any non-essential surgeries during April/2020. Still, most performed emergency cases, as recommended by the guidelines. In addition, it was interesting to observe urologists’ estimates of time beyond which delaying surgeries would harm patients, which seem to be in fair agreement with the expert recommendations.(34)

A clinical infection by the SARS-CoV-2 was reported by 13.5% of participants, including 5.2% with unequivocal laboratory confirmation. These numbers must be evaluated cautiously since they were self-reported. They seem to be much higher than expected, especially considering that the majority of the individuals did not participate in the COVID-19 front-line. A recent study showed that 5% of the population of the six more affected districts of the city of Sao Paulo had serological evidence of coronavirus infection.(35) Urologists are certainly more exposed than the general population since they visit hospitals and clinics where the risk of infection with COVID-19 is increased. What seems to be very worrisome is the high rates of COVID-9 requiring hospitalization, which was necessary for approximately 34% of the infected participants. Sadly, as we write this manuscript, we are aware of 4 urologists who passed away due to coronavirus infections. They all lived in states among those with the highest infection rates.

Since the start of Covid-19 a significant impact was observed in a number of health indicators of Brazilian urologists. Almost one third of the participants (32%) in our study reported being moderately healthy or not healthy. We believe that the mental afflictions resulting from the COVID pandemic may have an important role in this self-reported perception of general well-being although other factors such as changes in diet and physical activities may also play a role in this scenario.(36). The economic recession already in display represents an additional burden on mental health (37).

Although most reported their weight remained stable, 32.9% of the participants had some weight gain while 19.4% noticed weight loss. Physical activity was reduced for most urologists, while a small group reported an increase in physical activity. An increase in alcohol use was almost twice more common than the opposite, during the first two months of COVID-19 in Brazil. Finally, sexual activity was reported as stable by half of the participants but more than one third reported reduced sexual activity and only 13% reported an increase. Similar changes affecting the general population have been predicted by experts since the outbreak of the pandemic.(38–42) So far, however, few studies have been published evaluating such important issues. When it comes to physicians, we were not able to find any study reporting on the impact of COVID-19 in such parameters. A number of factors may be contributing to the reported changes in health parameters, including the stress associated with social distancing, fear of contamination and reduction of income. All may have a powerful effect on appetite and alcohol misuse and inhibit sexual activity.(39, 41, 42) Home confinement is also an obvious factors that might influence alcohol misuse, physical and sexual activity.(38, 40, 41, 43, 44) Our findings reinforce the need to implement interventions to promote health and wellbeing during the COVID-19 pandemic among health care workers. They should also include positive sexual health messages to help mitigate the detrimental effects of the pandemic.

We found it important to evaluate how Brazilian urologists are coping and occupying their free time during COVOID-19, especially considering the exceedingly long free time that most are having due to the drastic reduction of professional activities and the need to be confined at their homes. We found that the top activities occupying urologists’ time are use of social media (Facebook, Instagram, Tweeter), watching movies and TV shows, spending time with their families and physical activities. Other activities such as watch the news, study urology or COVID-19, play musical instruments, spirituality and the reading of non-medical books were also reported.

## 5. Conclusions

COVID-19 has introduced massive disturbances in the professional and personal lives of Brazilian urologists. The effects of the major cut-down in patient care, including patient visits and all kind of surgical procedures are still unknown, but will certainly be worsened with the prolongation of the pandemics. Also preoccupying are the distressing consequences of the pandemic on physicians’ health and personal lives. The uncertainties regarding the duration of the pandemic make this moment even more stressful for urologists and physicians alike, aggravating the perceptions of the occasion. Although this study only included urologists, the findings are probably applicable to most surgical and many clinical specialties that are not directly involved in caring for patients with COVID-19.

## Data Availability

The database is under the care of the Brazilian Society of Urology.

## Acknowledgments

The authors would like to thank the participants of the study for their time. Technical support was provided by Ricardo de Morais.

## Dedicatory

As we prepared this manuscript, we were aware of 4 urologists who died due to coronavirus infection: Aluízio Gonçalves da Fonseca; Fernando Jordão de Souza; Osmar Seabra; Pasquale Francisco Giglio. This work is dedicated to their memory and to all the health care workers who lost their lives to COVID-19.

## 9. SUPPLEMENT

List of Supplementary Materials

Supplementary material 1. Web-based survey with 39 questions, translated to English.

## REFERENCES

1. McFall JK. J; Morgan, L. A third of the global population is on coronavirus lockdown – here’s our constantly updated list of countries and restrictions. Business Insider Australia. 2020.

2. Dunford DD. B.; Stylianou, N.; Lowtherrr, E.; Ahmed, M.; Arenas, I. Coronavirus: The world in lockdown in maps and charts. https://www.bbccom/news/world-52103747. 2020.

3. Boletim Epidemiológico Especial. COE-COVID19. Secretaria de vigilância em saúde – Ministério da Saúde. https://www.saude.gov.br/images/pdf/2020/May/21/2020-05-19---BEE16---Boletim-do-COE-13h.pdf2020.

4. Engle J. How Is the Coronavirus Outbreak Affecting Your Life? The New York Times. 2020; https://www.nytimes.com/2020/03/20/learning/how-is-the-coronavirus-outbreak-affectingyour-life.html.

5. Mehrotra AC. M.; Linetsky, D.; Hatch, H.; Cutler, D. The Impact of the COVID-19 Pandemic on Outpatient Visits: A Rebound Emerges. The Commonwealth Fund. 2020; https://www.commonwealthfund.org/publications/2020/apr/impact-covid-19-outpatient-visits(Ateev Mehrotra, Michael Chernew, David Linetsky, Hilary Hatch, and David Cutler).

6. Margel D, Ber Y. Changes in Urology After the First Wave of the COVID-19 Pandemic. Eur Urol Focus. 2020.

7. Phe V, Karsenty G, Robert G, Game X, Cornu JN. Widespread Postponement of Functional Urology Cases During the COVID-19 Pandemic: Rationale, Potential Pitfalls, and Future Consequences. Eur Urol. 2020.

8. Nowroozi A, Amini E. Urology practice in the time of COVID-19. Urol J. 2020;17(3):326.

9. Ribal MJ, Cornford P, Briganti A, Knoll T, Gravas S, Babjuk M, et al. European Association of Urology Guidelines Office Rapid Reaction Group: An Organisation-wide Collaborative Effort to Adapt the European Association of Urology Guidelines Recommendations to the Coronavirus Disease 2019 Era. Eur Urol. 2020.

10. Gadzinski AJ, Ellimoottil C. Telehealth in urology after the COVID-19 pandemic. Nat Rev Urol. 2020.

11. Boehm K, Ziewers S, Brandt MP, Sparwasser P, Haack M, Willems F, et al. Telemedicine Online Visits in Urology During the COVID-19 Pandemic-Potential, Risk Factors, and Patients’ Perspective. Eur Urol. 2020.

12. Ferneini EM. The Financial Impact of COVID-19 on Our Practice. J Oral Maxillofac Surg. 2020.

13. Haleem A, Javaid M, Vaishya R. Effects of COVID 19 pandemic in daily life. Curr Med Res Pract. 2020.

14. Tull MT, Edmonds KA, Scamaldo KM, Richmond JR, Rose JP, Gratz KL. Psychological Outcomes Associated with Stay-at-Home Orders and the Perceived Impact of COVID-19 on Daily Life. Psychiatry Res. 2020;289:113098.

15. Stensland KD, Morgan TM, Moinzadeh A, Lee CT, Briganti A, Catto JWF, et al. Considerations in the Triage of Urologic Surgeries During the COVID-19 Pandemic. Eur Urol. 2020;77(6):663–6.

16. Ficarra V, Novara G, Abrate A, Bartoletti R, Crestani A, De Nunzio C, et al. Urology practice during COVID-19 pandemic. Minerva Urol Nefrol. 2020.

17. Carneiro A, Wroclawski ML, Nahar B, Soares A, Cardoso AP, Kim NJ, et al. Impact of the COVID-19 Pandemic on the Urologist’s clinical practice in Brazil: a management guideline proposal for low-and middle-income countries during the crisis period. Int Braz J Urol. 2020;46(4):501–10.

18. Carter MR, Aaron E, Nassau T, Brady KA. Knowledge, Attitudes, and PrEP Prescribing Practices of Health Care Providers in Philadelphia, PA. J Prim Care Community Health. 2019;10:2150132719878526.

19. Garcia AV, Jeyaraju M, Ladd MR, Jelin EB, Bembea MM, Alaish S, et al. Survey of the American Pediatric Surgical Association on cannulation practices in pediatric ECMO. J Pediatr Surg. 2018;53(9):1843–8.

20. Shanthanna H, Moisuik P, O’Hare T, Srinathan S, Finley C, Paul J, et al. Survey of Postoperative Regional Analgesia for Thoracoscopic Surgeries in Canada. J Cardiothorac Vasc Anesth. 2018;32(4):1750–5.

21. Lavallee LT, Fergusson D, Mallick R, Grenon R, Morgan SC, Momoli F, et al. Radiotherapy after radical prostatectomy: treatment recommendations differ between urologists and radiation oncologists. PLoS One. 2013;8(11):e79773.

22. Plata M, Bravo-Balado A, Robledo D, Castano JC, Averbeck MA, Plata MA, et al. Trends in pelvic organ prolapse management in Latin America. Neurourol Urodyn. 2018;37(3):1039–45.

23. Important and urgent – next steps on NHS response to COVID-19. NHS England. 2020; https://www.england.nhs.uk/coronavirus/wp-content/uploads/sites/52/2020/03/urgent-nextsteps-on-nhs-response-to-covid-19-letter-simon-stevens.pdf.

24. COVID-19: recommendations for management of elective surgical procedures. American College of Surgeons. 2020.

25. Mangialardi ML, Orrico M, Mangialardi N. Routine in an Italian high-volume Vascular Surgery Unit during the COVID-19 era: how the pandemic changed the vascular daily practice. Ann Vasc Surg. 2020.

26. Morlacco A, Motterle G, Zattoni F. The multifaceted long-term effects of the COVID-19 pandemic on urology. Nat Rev Urol. 2020.

27. Amparore D, Claps F, Cacciamani GE, Esperto F, Fiori C, Liguori G, et al. Impact of the COVID-19 pandemic on urology residency training in Italy. Minerva Urol Nefrol. 2020.

28. Hussain K, Dewan V, Ali T, Al Shakarchi J. The impact of the COVID-19 pandemic on the provision of surgical care. J Surg Case Rep. 2020;2020(4):rjaa087.

29. Shah JP. The impact of COVID-19 on Head and Neck surgery, education, and training. Head Neck. 2020.

30. Ministério da Saúde. Portaria número 467, de 20 de marcço de 2020. Diário Oficial da União. 2020; http://www.in.gov.br/en/web/dou/-/portaria-n-467-de-20-de-marco-de-2020-249312996.

31. Portnoy JM, Wu AC. Is Telemedicine as Effective as Usual Care? J Allergy Clin Immunol Pract. 2019;7(1):217–8.

32. Orlando JF, Beard M, Kumar S. Systematic review of patient and caregivers’ satisfaction with telehealth videoconferencing as a mode of service delivery in managing patients’ health. PLoS One. 2019;14(8):e0221848.

33. Burki TK. Cancer guidelines during the COVID-19 pandemic. Lancet Oncol. 2020;21(5):629–30.

34. Heldwein FL, Loeb S, Wroclawski ML, et al. A systematic review on guidelines and recommendations for urology standard of care during COVID-19 pandemic. Eur Urol Focus. 2020;in press.

35. Galhardo RB. D. Estudo inédito detecta anticorpos ao coronavírus em 5% dos moradores da cidade de São Paulo. O Estado de São Paulo. 2020; https://saude.estadao.com.br/noticias/geral,estudo-inedito-detecta-anticorpos-aocoronavirus-em-5-dos-moradores-da-cidade-de-sao-paulo,70003304706.

36. Pfefferbaum B, North CS. Mental Health and the Covid-19 Pandemic. N Engl J Med. 2020.

37. Frasquilho D, Matos MG, Salonna F, Guerreiro D, Storti CC, Gaspar T, et al. Mental health outcomes in times of economic recession: a systematic literature review. BMC Public Health. 2016;16:115.

38. Pecanha T, Goessler KF, Roschel H, Gualano B. Social isolation during the COVID-19 pandemic can increase physical inactivity and the global burden of cardiovascular disease. Am J Physiol Heart Circ Physiol. 2020;318(6):H1441–H6.

39. Bhutani S, Cooper JA. COVID-19 related home confinement in adults: weight gain risks and opportunities. Obesity (Silver Spring). 2020.

40. Jacob L, Smith L, Butler L, Barnett Y, Grabovac I, McDermott D, et al. COVID-19 social distancing and sexual activity in a sample of the British Public. J Sex Med. 2020.

41. Clay JM, Parker MO. Alcohol use and misuse during the COVID-19 pandemic: a potential public health crisis? Lancet Public Health. 2020;5(5):e259.

42. Robinson BE. What Is “Quarantine 15”? Psychology Today. 2020; https://www.psychologytoday.com/us/blog/the-right-mindset/202003/what-is-quarantine-15.

43. Riley T, Sully E, Ahmed Z, Biddlecom A. Estimates of the Potential Impact of the COVID-19 Pandemic on Sexual and Reproductive Health In Low-and Middle-Income Countries. Int Perspect Sex Reprod Health. 2020;46:73–6.

44. Yip PSF, Chau PH. Physical Distancing and Emotional Closeness Amidst COVID-19. Crisis. 2020;41(3):153–5.

